# Survival risk heterogeneity among patients with NSCLC receiving nivolumab visualized by risk scores generated from deep learning method DeepSurv using tumor gene mutations

**DOI:** 10.64898/2026.02.15.26346303

**Authors:** Yoshiaki Nishiyama, Nobuaki Nishiyama

## Abstract

Immunotherapy with immune checkpoint inhibitors and immunotherapy combined with chemotherapy have represented promising treatments for NSCLC patients leading to prolonged survival. However, the majority of patients with advanced NSCLC have a poor prognosis. The identification and development of biomarkers for stratifying responders and non responders to immune checkpoint inhibitors contribute to unravel the mechanism of immune checkpoint pathway and the immune tumor interaction underlying the responses and are urgently needed to improve clinical outcomes of immune checkpoint inhibitor treatment. In this study, we analyzed the clinical and gene mutation data of NCSLC patients treated with nivolumab containing immunotherapy or nivolumab containing immunotherapy combined with chemotherapy (the immunotherapy treated group, n=119) and chemotherapy alone (the chemotherapy alone treated group, n=991) extracted from the MSK CHORD dataset. A DeevSurv model, a deep learning based extension of the Cox proportional hazards model was trained to generate survival risk score of each patient with binary statuses of thirty one gene mutations as input features into the model. The thirty one genes were selected based on population level mutation frequency, patient level variance in mutation status, and univariate Cox proportional hazards analyses evaluating the association between the presence or absence of each gene mutation and overall survival. The performance of the trained DeepSurv model was evaluated on the test set of the immunotherapy treated group using the concordance indexes (C index). The trained model was subsequently applied without retraining to the entire chemotherapy alone treated group as a control. The resulting C indexes for the immunotherapy treated group and chemotherapy alone treated group were 0.789 and 0.483, respectively. All patients within each group were divided into high and low risk groups according to the median predicted risk score. Kaplan Meier survival curves of high and low risk groups (n=43 vs n=70) in the immunotherapy treated group revealed a significant separation (log rank p<0.001), whereas no separation was observed in chemotherapy alone treated group (p=0.62). In the combined cohort of the immunotherapy treated group and chemotherapy alone treated group, the interaction between the DeepSurv derived risk score and treatment modality was significant (HR for interaction 1.47, 95% CI from 1.32 to 1.65, p<0.005), suggesting the DeepSurv derived risk score predictive value specific to the immunotherapy. Principal component analysis and permutation importance analysis were performed as complementary analyses to assess individual genes associated with the DeepSurv derived risk score and identified ZFHX3, SMARCA4, ALK, BTK, and NOTCH2 as major contributors to survival risk stratification. Collectively. we suggested that nonlinear coupling pattern of 31 tumor gene mutation statuses in the DeepSurv model captures the heterogeneity of survival risk among nivolumab containing immunotherapy or nivolumab containing immunotherapy combined with chemotherapy treated patients with NSCLC which was visualized as clear separation between high risk and low risk groups divided by the median value of the risk scores.

## 1. Introduction

Immunotherapy with immune checkpoint inhibitors such as anti-PD-1, anti-PD-L1 and anti-CTLA4 agents and immunotherapy combined with chemotherapy have represented promising treatments of choice for patients with NSCLC based on significant prolonged survival compared with chemotherapy alone[1-5]. However, the majority of patients with advanced NSCLC have a routinely poor prognosis. The identification and development of biomarkers for stratifying responders and non-responders to immune checkpoint inhibitors contribute to unravel the mechanism of immune checkpoint pathway and the immune–tumor interaction underlying the responses and are urgently needed to improve clinical outcomes of immune checkpoint inhibitor treatments [6].

While the association between high expression of tumor PD-L1 and clinical benefits from immune checkpoint inhibitors in patients with NSCLC has been evidenced by clinical trials and real-world data across various treatment strategies including immune checkpoint inhibitor monotherapy, combinations of multiple immune checkpoint inhibitors, and combinations with chemotherapy, there are accumulating evidences that the treatments with immune checkpoint inhibitors represent clinical benefit in patients with PD-L1 expression level less than 1% [7-13]. The association between higher tumor mutation burden (TMB) and durable response to immune checkpoint inhibitors in NSCLC has been also observed [14.15]. However, overall survival benefit from immune checkpoint inhibitor treatments in patients with NSCLC were frequently found regardless of TMB [16-18], leading to integrated biomarkers of TMB combined with tumor PD-L1 expression [19] or gene alterations of STK11, KEAP1, and EGFR [20]. These efforts towards refined biomarker development suggest the selection of multiple tumor gene mutations that influence the clinical outcomes of immune checkpoint inhibitor treatment in NSCLC, which may better capture underlying mechanisms of the immune checkpoint pathway and the immune–tumor interaction.

In line with this direction, recent machine learning–based studies have attempted to identify mutation-derived genomic signatures that predict immunotherapy benefit in NSCLC. Liu et al. classified the patients with NSCLC receiving immune checkpoint inhibitor treatments into therapeutic benefit and non-benefit groups based on progression-free survival duration and applied the XGBoost algorithm to evaluate the importance of each mutated gene for distinguishing treatment benefit from treatment non-benefit patients [21]. XGBoost constructs an ensemble of decision trees and gene mutation features are repeatedly ranked using permutation-based importance across 350 iterations. The top-ranking genes converged to a stable set of 88 mutated genes. In this response-label driven machine learning method with external validation, the resulting 88 mutated gene signature showed superior predictive performance compared with tumor mutational burden for identifying patients who benefit from immune checkpoint inhibitor treatment in NSCLC[21]. Fomin et al. applied a genetic algorithm (GA) coupled with a Bernoulli Naive Bayes classifier to identify multiple gene mutations in NSCLC, using two patient outcome labels, durable clinical benefit and innate resistance as supervisory signals[22]. In this approach, GA proposes a mutated gene subset from hundreds of somatic mutations encoded as binary features and each feature subset is evaluated by the Bernoulli Naive Bayes classifier trained on the two patient outcome labels. The classifier’s cross-validated accuracy is then returned to the GA as the fitness score, resulting in increasingly predictive mutated gene subsets for immune checkpoint inhibitor response. Through this response-label driven method, the study ultimately identified 33 genomic signatures with strong predictive capacity for immune checkpoint inhibitor response in NSCLC[22].

In the present study, we aimed to develop a deep learning model, DeepSurv [23] with tumor multiple gene mutation statuses in NSCLC as input features to capture non-linear patterns of gene mutation combinations underlying survival heterogeneity among patients receiving nivolumab-containing immunotherapy or nivolumab-containing immunotherapy combined with chemotherapy. Using overall survival time and censoring status as the supervisory signals, the DeepSurv model which is a deep learning– based extension of the Cox proportional hazards (Cox PH) model outputs an individualized continuous survival risk score for each patient, rather than the classification into two response groups in prior studies [21,22]. We selected 31 genes as input features for DeepSurv, based on population-level mutation frequency, patient-level variance in mutation status, and univariate Cox proportional hazards analyses evaluating the association between the presence or absence of each gene mutation and overall survival. The resulting risk scores suggested the survival heterogeneity among nivolumab treated patients, which was visualized as clear separation between high-risk and low-risk groups divided by the median value of the risk scores.

## 2. Materials and Methods

### 2.1. Clinical and genomic data sources

Clinical and genomic data were obtained from the large-scale structured oncologic dataset (MSK-CHORD) which has been provided as a public resource for real-world oncologic research[24]. Clinical and sample data were a single or multiple data collected along the treatment timeline for a patient. Only the data including a single or multiple sample data originated from primary NSCLC and with the status of tumor PD-L1 expression (positive, negative) were extracted. The resulting data were classified into the immunotherapy treated group and the chemotherapy-only treated group based on the information of administered agents such as nivolumab, pembrolizumab, cisplatin. In the immunotherapy treated group, the patients with a single sample were defined as having been with treated with nivolumab and the patients with multiple samples defined as having been treated with nivolumab at least once and other immune checkpoint inhibitors or chemethrerapeutic agents. In the chemotherapy alone treated group, the patients with a single or multiple samples were defined as having been treated with chemotherapeutic agents only, without any administration of immunotherapeutic agents. In the case of multiple samples for a patient, the most recent treatment data was selected. As a result, each clinical data described above corresponded to a patient with overall survival variables (OS_MONTHS, OS_STATUS). Somatic gene mutation data were obtained in a binary format indicating the presence or absence of each mutation for individual patients. Each feature correspondeds to a specific gene–alteration type (single nucleotide variant or structural variant).

### 2.2. Gene mutation feature selection

Genes with mutation status encoded in a binary format were selected stepwise as described below and the selected genes were used as input features to the DeepSurv model. Mutated genes with population-level mutation frequency of less than 1% within the immunotherapy treated group were excluded and further removal of genes with population-level mutation frequency in fewer than 3% or more than 30% of patients were performed, as such genes provide limited discriminatory information. Additionally, genes with minimal variance of mutation across patients in the immunotherapy treated group (variance ≤ 0.01) were removed to ensure sufficient heterogeneity for model learning. Univariate Cox proportional hazards analyses were applied to each of the remaining genes to evaluate the association between the presence or absence of gene mutation and overall survival. A relatively permissive significance threshold (p < 0.30) was used to avoid the excessive exclusion of potentially useful prognostic factor candidates. This multistep filtering process resulted in the selection of 31 gene mutation features.

### 2.3. DeepSurv training and evaluation

A feed-forward neural network model was constructed based on DeepSurv framework [23], which is an extension form of the Cox proportional hazards model. Using overall survival time and censoring status as the supervision, the model outputs an individualized continuous survival risk score for each patient. The input features for the DeepSurv model were 31 binary gene mutation variables as described above. The neural network architecture has fully connected hidden layers with rectified linear unit (ReLU) activation functions, including two hidden layers with 32 and 16 neurons, respectively. The immunotherapy treated group was randomly split into training and testing sets in a 70:30 ratio using stratified sampling with respect to censoring status. The 31 binary gene mutation input features were standardized using z-score normalization based on statistics derived from the training set and subsequently applied to the test set. Model training was performed using the Adam optimizer with a fixed learning rate of 5 × 10^−4^, a batch size of 16, and a maximum of 250 training epochs. Baseline hazards were estimated after training using the Breslow method as implemented in the pycox framework. Model performance was evaluated on the held-out test set using the concordance index (C-index). The trained DeepSurv model was subsequently applied without retraining to the entire chemotherapy alone treated group as a control and predictive performance was again quantified using the C-index. To visualize the degree of survival risk heterogeneity within each treated group, all patients within each group were divided into high- and low-risk groups according to the median predicted risk score, and Kaplan–Meier survival curves of high- and low-risk groups in each treated group were generated and compared with the log-rank test.

### 2.4. Cox proportional hazards regression analysis for prognostic and treatment effects

For the immunotherapy treated group, a Cox proportional hazards regression model with the interaction term between the DeepSurv-derived continuous risk score and tumor PD-L1 expression status was used to assess their association with overall survival. Additionally, a Cox proportional hazards regression model including a DeepSurv-derived risk score–by–treatment interaction term was used to assess whether the risk score has treatment (immunotherapy and chemotherapy alone) dependent association with overall survival. The trained DeepSurv model with the training set of the immunotherapy treated group was applied without retraining to all patients in both the chemotherapy alone treated and the immunotherapy treated group to calculate DeepSurv-derived risk scores. The resulting DeepSurv risk scores were used in the Cox regression model with an interaction term. These risk scores were centered using the mean value of all patients. The model includes two variables of the centered DeepSurv-derived risk score X, the treatment indicator (immunotherapy =1, chemotherapy-only=0) T, and the interaction X・T with the corresponding regression coefficients, β1, β2, and β3, respectively. β1 represents prognostic association of the DeepSurv-derived risk score with overall survival in chemotherapy alone treated patients (T=0), as both the treatment term and the interaction term equal zero. β2 represents the difference in overall survival between the two treatment groups at the mean value of the risk score (X=0) with the interaction term equals zero. β3 represents the extent which the association between the risk score and overall survival differs between the two treatments. This analysis allows a direct assessment of whether the DeepSurv-derived risk score functions as a treatment-dependent predictive factor for immunotherapy.

### 2.5. Principal component analysis of gene mutation profile within immunotherapy treated group

We hypothesized that survival risk heterogeneity within immunotherapy treated group could be captured by nonlinear coupling of the mutation status of 31 genes in the DeepSurv model. In contrast, to examine whether similar survival risk heterogeneity could be captured through linear combinations of the same gene mutation features, we performed principal component analysis (PCA). The 31 gene mutation features were standardized by centering each gene’s mutation values around its average across patients to remove differences in mutation frequency between genes and by rescaling each gene’s mutation values based on the degree of variation among patients to ensure that genes with very different mutation frequencies contribute comparably to the analysis. In the analysis of PCA, only 31 gene mutation statuses of patients within the immunotherapy treated group were used but not DeepSurv-derived risk scores and survival outcomes. For the visualization of analysis result, patients were annotated post hoc according to DeepSurv-defined risk groups (high-risk, low-risk) to assess whether PCA could reproduce the separation between the two risk groups in the plane of the first two principal components (PC1 and PC2). The gene mutation profile compressed into PC1-PC2 plane indicates the property inherent in the immunotherapy treated group. To obtain the insight about gene mutation specific to the immunotherapy treated group, loadings of individual gene mutations on PC1 were examined.

### 2.6. Permutation importance analysis of gene mutation features for the DeepSurv model

While survival risk heterogeneity within the immunotherapy treated group captured by the DeepSurv model is hypothesized to be due to nonlinear coupling of 31 gene mutation features, we examined how each gene mutation contributes to detect the survival risk heterogeneity by using permutation importance analysis on the held-out immunotherapy test set. The binary status of each gene mutation was randomly permuted among patients within immunotherapy test set, while the binary status of all other genes, survival times and censoring information of patients in the test set were kept unchanged. In detail, the column corresponding to a given gene in the feature matrix was randomly reordered across patients, then the original patient–gene association was disrupted while preserving the overall mutation frequency of that gene in the test set. The trained DeepSurv model was then applied to the permuted test set without retraining, and survival risk scores were recalculated. This permutation procedure was repeated multiple times for each gene, and the importance of each gene was quantified as the mean decrease in C-index across permutations with standard deviation. The resulting change of C-index indicate the importance of the targeted gene for capturing survival risk heterogeneity within the immunotherapy treated group and provides insights into biological functions underlying immunotherapy benefits.

### 2.7. Statistical analysis and software

Analyses were performed using Python (version 3.11.13) via the Spyder IDE (Anaconda distribution), employing pandas (version 2.3.1), NumPy (version 1.26.4), scikit-learn (version 1.7.1) and the lifelines library (version 0.30.0). The DeepSurv model was implemented with PyTorch (version 2.5.1) in conjunction with the pycox library (version 0.3.0). All figures were created using matplotlib (version 3.10.0).

## 3. Results

Within the immunotherapy treated group (n=119), survival curves depending on PD-L1 expression were estimated by using univariate Kaplan–Meier method as shown in Figure 1. The median overall survival of patients with PD-L1 positive tumor was longer compared with that of patients with PD-L1 negative tumor (36.6 vs. 19.4 months), but the difference was not significant (log-rank test: χ^2^ = 0.48, p = 0.49).

**Figure 1.**
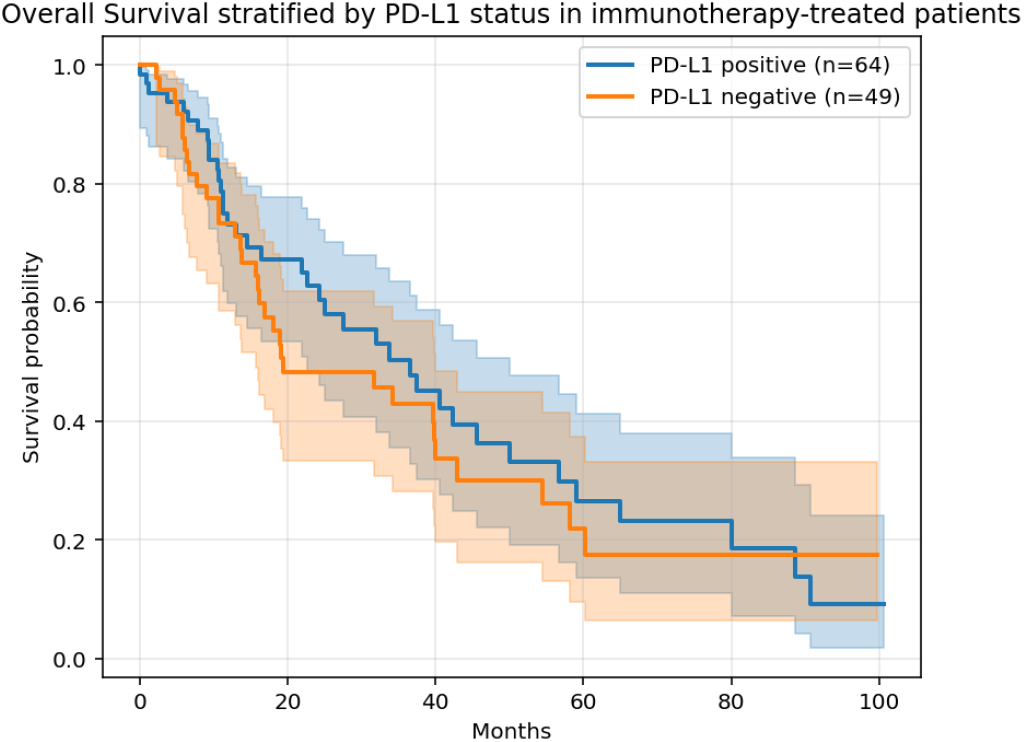
Kaplan–Meier overall survival curves corresponding to patients with positive PD-L1 expression and patients with negative PD-L1 expression within the immunotherapy treated group. The shaded areas indicate 95% confidence intervals.

While the observed trend toward improved survival in the immunotherapy group limited to patients treated with agents including nivolumab is consistent with survival curves obtained in the immunotherapy treated NSCLC cohort of the original MSK-CHORD [24], PD-L1 expression was significantly associated with a hazard ratio of approximately 0.6 for overall survival in the large-scale MSK-CHORD study, suggesting that tumor PD-L1 expression alone was insufficient to stratify overall survival within our immunotherapy treated group with limited number of patients.

By using 31 gene mutation status as input features described in Method Section 2.2, the DeepSurv model was trained on the train set (70%) of the immunotherapy treated group, and model performance was evaluated on the test set (30%) using the concordance indexes (C-index). The trained DeepSurv model was subsequently applied without retraining to the entire chemotherapy alone treated group as a control. The resulting C-index values were 0.789 and 0.483, respectively. Using the trained DeepSurv model, risk scores were calculated for the entire immunotherapy-treated group and the entire chemotherapy-only treated group. All patients within each group were divided into high- and low-risk groups according to the median predicted risk score. Kaplan–Meier survival curves of high- and low-risk groups (n=43 vs n=70) in the immunotherapy treated group are shown in Figure 2a and reveals a pronounced separation between the high- and low-risk groups, with significantly worse overall survival in the high-risk group (log-rank p << 0.001). Kaplan–Meier survival curves of high- and low-risk groups (n=265 vs n=726) are shown in Figure 2b and no significant survival difference was observed between the risk groups (log-rank p = 0.62). These findings suggest that the DeepSurv-derived risk score based on 31 gene mutation status captures survival heterogeneity specific to the immunotherapy treated group.

**Figure 2.**
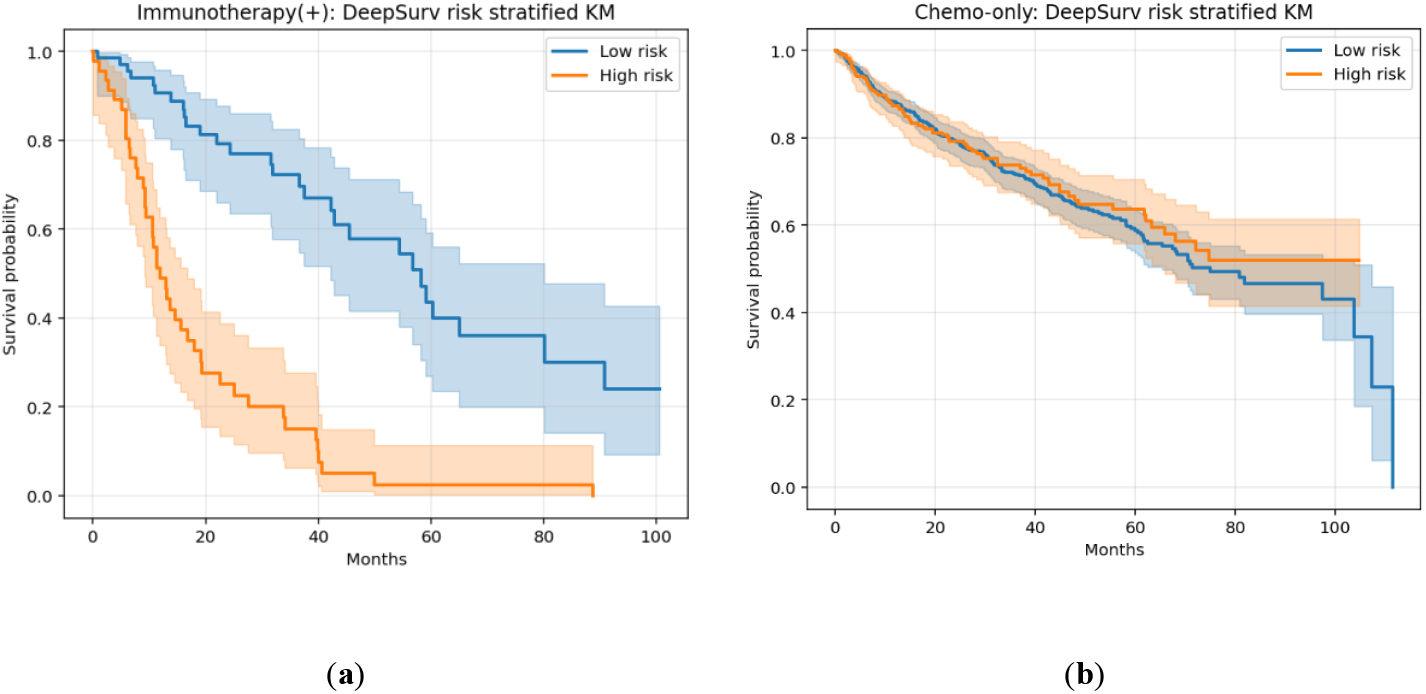
All patients within each group were divided into high- and low-risk groups according to the median of the DeepSurv-derived risk scores. a)Kaplan–Meier curves in the immunotherapy treated group (high risk vs low risk; n=43 vs 70) show significant survival separation (C-index=0.789; log-rank p<0.001). b) Kaplan–Meier curves stratified by the same DeepSurv risk score in the chemotherapy alone treated group (high risk vs low risk; n=265 vs 726) show no significant survival difference (C-index=0.483; log-rank p=0.62).

For the immunotherapy treated group, a multivariable Cox regression model with the interaction term between the DeepSurv-derived continuous risk and tumor PD-L1 expression status was used to assess their association with overall survival. The DeepSurv-derived risk score was significantly associated with overall survival and this association was similar for both tumor PD-L1 nagative and PD-L1 positive status (HR = 1.80, 95% CI 1.49–2.18 for negative status vs HR = 1.53, 95% CI 1.16–2.03 for positive status). Tumor PD-L1 expression status was not significantly associated with overall survival (HR = 0.79, 95% CI 0.47– 1.31, p = 0.36) with adjustment for the DeepSurv-derived risk score. The interaction between the DeepSurv-derived risk score and tumor PD-L1 expression status was not also significant (HR = 0.85, 95% CI 0.69-1.04, p=0.11). These findings indicate that the DeepSurv-derived risk score is the main contributor to highly predictive performance (C-index = 0.80, likelihood ratio test p<0.001) of the Cox regression model and are consistent with no significant difference in overall survival between patients with PD-L1 positive and PD-L1 negative tumor within the immunotherapy-treated group as shown in Figure 1.

For all patients by combining immunotherapy treated patients (n=113) and chemotherapy alone treated patients (n=991), a multivariable Cox proportional hazards model was applied to assess whether the DeepSurv-derived risk score was associated with overall survival depending on treatment modality. The model included the centered DeepSurv-derived risk score X, a treatment indicator (T; immunotherapy = 1, chemotherapy-only = 0), their interaction X・T and the corresponding regression coefficient β_1_, β_2_, and β_3_, respectively. The coefficient β_1_ was not significant (HR = 0.99, 95% CI 0.95–1.04, p = 0.84) for the association of the centered DeepSurv-derived risk score and overall survival, indicating that the risk score has little prognostic relevance in chemotherapy alone treated patients. The coefficient β_2_ represented a significant difference between the immunotherapy treated group and the chemotherapy alone treated group at the mean value (X=0) of the risk score (HR = 2.51, 95% CI 1.89–3.35, p < 0.005) and the hazard ratio indicates the estimated hazard of death was more than twice as high in the immunotherapy treated group compared with the chemotherapy alone treated group. This higher hazard ratio may reflect poorer baseline prognostic profiles of immunotherapy treated group compared with those of chemotherapy alone treated group originating from real-world, non-randomized dataset, consistent with Kaplan–Meier survival curves shown in Figure 2a and 2b. The coefficient β_3_ which represents the difference of the association between the risk score and overall survival by treatment modality was significant (HR = 1.47, 95% CI 1.32–1.65, p < 0.005) and the high hazard ratio for the interaction term indicates that the effect of the risk score on mortality is enhanced in the immunotherapy treated group compared with the chemotherapy alone treated group. Together with the absence of a significant association between the risk score and overall survival in the chemotherapy alone treated group, these findings suggest that the DeepSurv-derived risk score based on nonlinear coupling of 31 gene mutation statuses may serve as an immunotherapy-specific predictive biomarker.

To assess how the intrinsic patterns of 31 selected gene mutation statuses within the immunotherapy treated group are captured with a linear combination method of features, principal component analysis (PCA) was performed.

The PCA did not use the DeepSurv-derived risk scores and survival outcomes and the compressed patterns of 31 gene mutation status into PC1 - PC2 plane is therefore independent on survival risk. The result is shown in Figure 3, in which each patient was classified post hoc according to the DeepSurv-derived high risk group and low risk group as shown in Figure 2a. The two DeepSurv-derived risk groups have substantial overlap along the PC1 axis, suggesting that linear combinations of gene mutation features are insufficient to capture the survival risk heterogeneity visualized by the DeepSurv-derived risk scores. PC1 loading which indicates how strongly each gene contributes the main axis of mutational variation revealed that several genes including PIK3C2G, NOTCH2, BTK, NSD1, IL-7R, SMARCA4, and EPHA7 have large influences as shown in Figure 4. These identified gene mutations are likely to characterize intrinsic properties of tumors within the immunotherapy treated group, rather than mutations directly responsible for the observed heterogeneity through the DeepSurv-derived survival risk.

**Figure 3.**
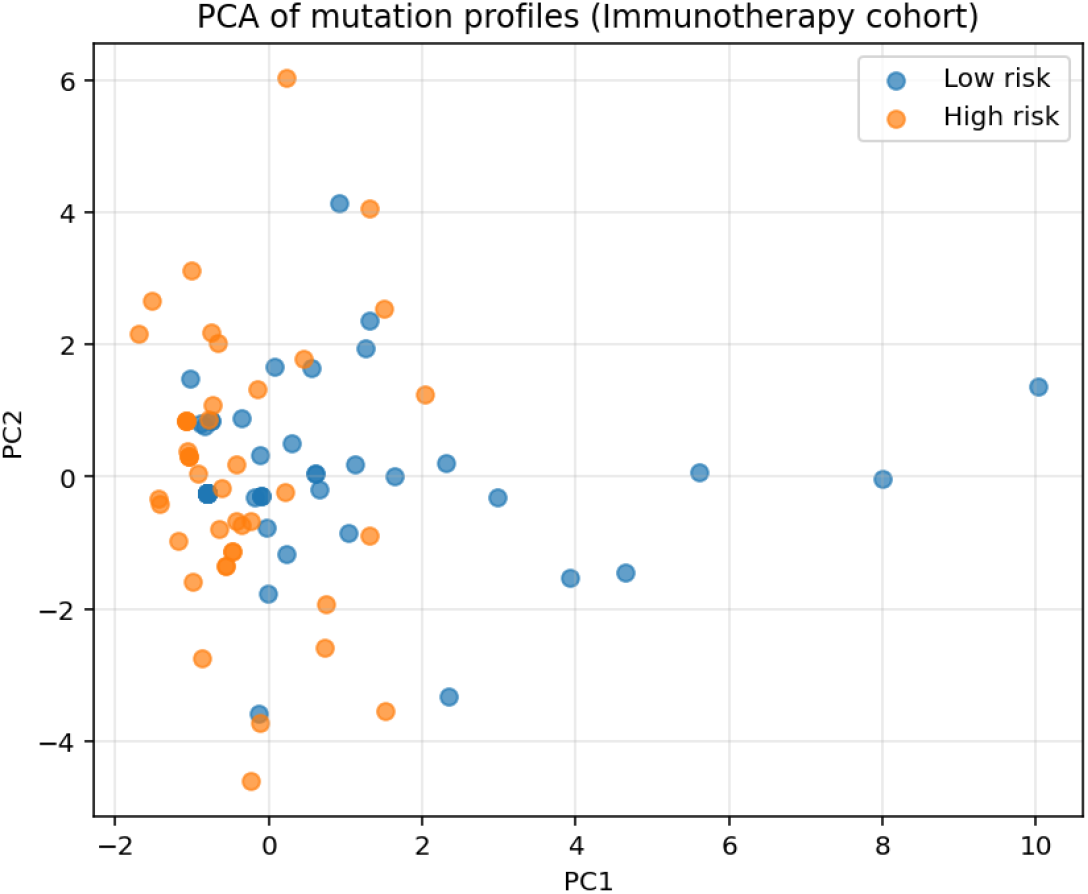
PCA of 31 gene mutation profiles compressed into the PC1-PC2 plane in the immunotherapy treated group. The PCA did not use the DeepSurv-derived risk scores and survival outcomes and post hoc labeling by DeepSurv risk groups (high risk and low risk) shows a trend toward separation between DeepSurv-derived high risk group and low risk group.

**Figure 4.**
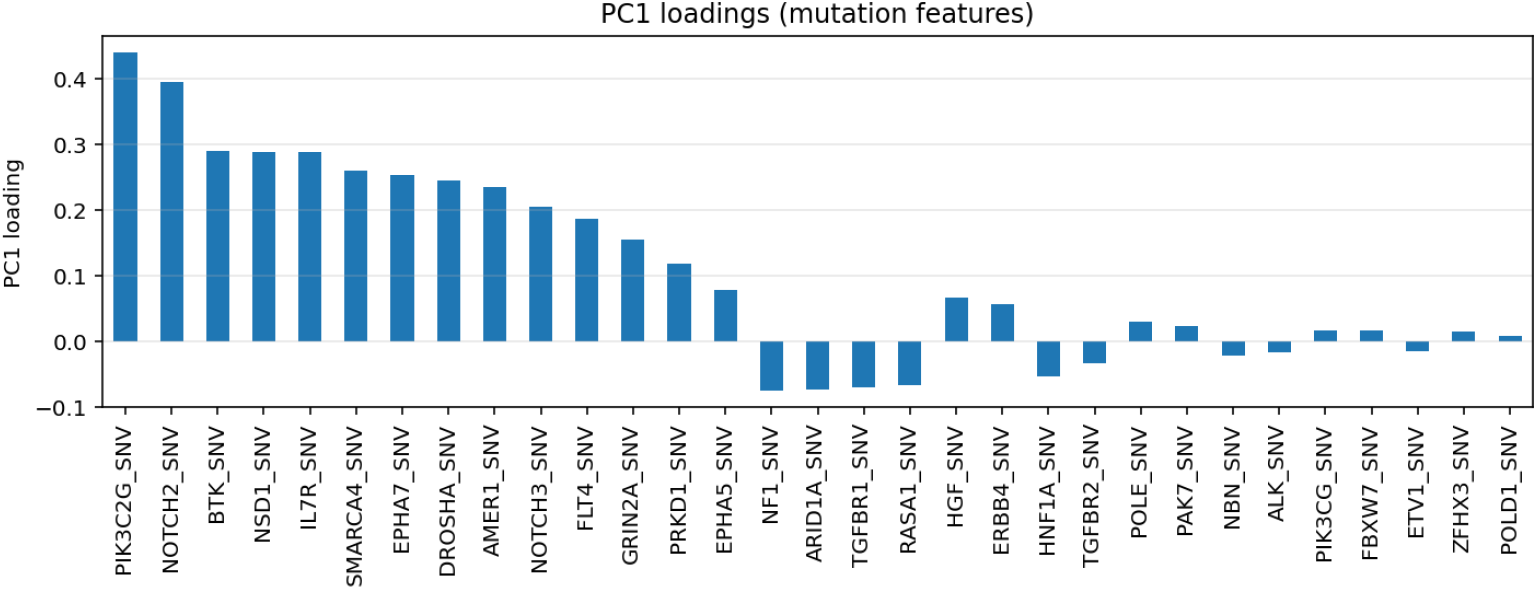
PC1 loading which indicates how strongly each gene contributes the main axis of mutational variation revealed that several genes including PIK3C2G, NOTCH2, BTK, NSD1, IL-7R, SMARCA4, and EPHA7 have large influences.

While survival risk heterogeneity within the immunotherapy treated group captured by the DeepSurv model is hypothesized to be due to nonlinear coupling of 31 gene mutation features, we examined how each gene mutation contributes to detect the survival risk heterogeneity by using permutation importance analysis on the held-out immunotherapy test set. As shown in Figure 5, repeated permutation of individual gene mutation features resulted in marked decreases in C-index for several genes, prominently ZFHX3, SMARCA4, ALK, BTK, and NOTCH2, suggesting that regulatory networks including these genes are potentially associated with survival risk of patients treated with immune checkpoint inhibitors including nivolumab.

**Figure 5.**
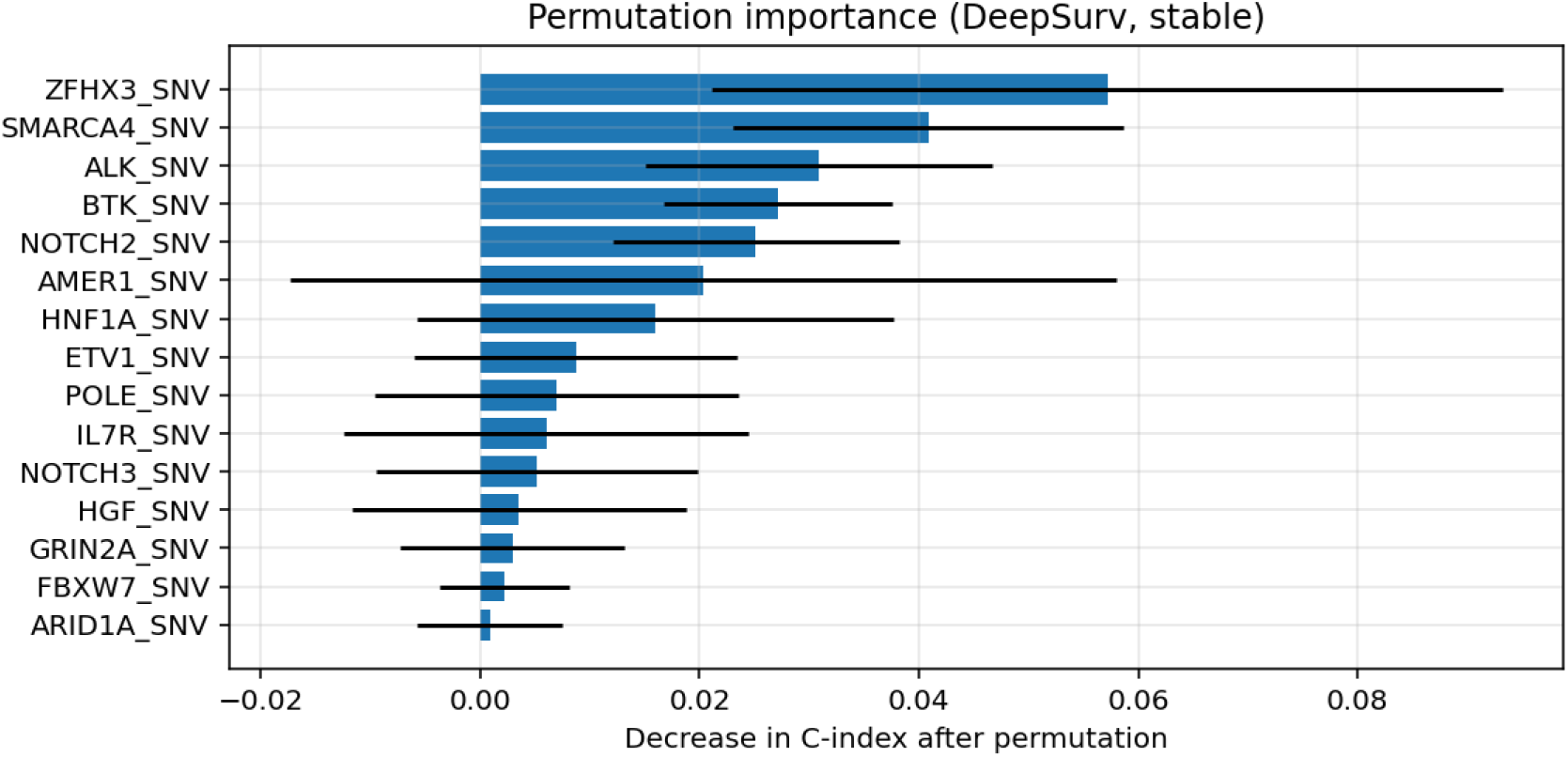
Permutation importance of each gene was obtained as the mean decrease in C-index across repeated permutations in the held-out immunotherapy test set. Error bars indicate standard deviation.

## 4. Discussion

In this study, we suggested that non-linear coupling pattern of 31 tumor gene mutation statuses in a DeepSurv model captures the heterogeneity of survival risk among NSCLC patients treated with nivolumab-containing immunotherapy or nivolumab-containing immunotherapy combined with chemotherapy, that is, among the immunotherapy treated group and fails to identify a heterogeneity of survival risk within the chemotherapy alone treated group. With a multivariable Cox proportional hazards model, the significant interaction between the DeepSurv-derived risk score and treatment modality (the immunotherapy treated group and the chemotherapy alone treated group) further supports the interpretation that the nonlinear coupling pattern of thirty-one tumor gene mutation statuses functions as an immunotherapy-specific predictive biomarker for NSCLC patients. However, the biological interpretability of the nonlinear interactions learned by DeepSurv, that is, how regulatory networks involving these 31 mutated or wild genes are underlying in survival risk or benefit within the immuotherapy-treated patients with NSCLC is inherently limited and difficult to elucidate directly. To address this, we further performed PCA and permutation-based analysis as complementary analyses to identify individual genes associated with the DeepSurv-derived risk score. PCA approximates each patient’s 31 gene mutation profile using linear combinations of the 31 binary features. PC1 and PC2 represent weighted linear combinations of these features and the loadings of each gene on PC1 and PC2 indicate the extent to which each gene contributes to the principal linear patterns that approximate the non-linear and biological complex interactions of the 31 genes as shown in Figure 4. In addition, we identified which individual gene mutation status of the 31 genes contribute to detect the DeepSurv-derived survival risk heterogeneity by using permutation importance analysis as shown in Figure 5.

The 31 genes were identified using population-level frequency thresholds, variance filtering, and univariate Cox analyses, without using predefined treatment-response labels in the preceding studies [21,22]. The identified 31 genes were ALK, EPHA5, EPHA7, ERBB4, FLT4, HGF, IL7R, NF1, RASA1, PIK3C2G, PIK3CG, and BTK, NOTCH2, NOTCH3, TGFBR1, TGFBR2, POLE, POLD1, NBN, DROSHA, and GRIN2A, ARID1A, SMARCA4, NSD1, FBXW7, ZFHX3, HNF1A, AMER1, suggesting these genes involved in distributed and interconnected regulatory networks [25-40]. The genes identified with high PC1 loading such as PIK3C2G are likely to represent intrinsic tumor characteristics varying highly among patients treated with immunotherapy, whereas the genes with high permutation importance such as ZFHX3 are likely to be functionally relevant to survival risk under immunotherapy. BTK, NOTCH2, and SMARCA4 highly ranked in both PC1 loading and permutation importance were suggested to be critical determinants of survival risk depending on their mutation status and the interpretation is partially consistent with the trend toward the separation between DeepSurv-derived high risk group and low risk group, although the separation along the PC1 axis was imperfect with substantial overlap as shown in Figure A recent study identified the mutation status of PIK3C2G as a prognostic factor significantly associated with overall survival in patients with stage IIb and IIIa lung adenocarcinoma [41], suggesting that the mutation status of PIK3C2G underlies survival heterogeneity among NSCLC patients and is consistent with being PIK3C2G top-ranked in the PC1 loading in our study. Formin et al. identified PIK3C2G as one of gene mutation signatures associated with immune checkpoint inhibitor responders in NSCLC and demonstrated that it represents one of immunotherapy specific predictive factors compared with chemotherapy [22], consistent with our result showing that PIK3C2G is included among 31 gene mutation statuses with non-linear coupling in the present DeepSurv model, although it is not ranked in the permutation importance analysis. Studies analyzing the relationship between gene mutations and the efficacy of immune checkpoint inhibitor treatments in NSCLC patients demonstrated that mutations in ZFHX3 [42] and NOTCH1/2/3 [43] are significantly associated with longer overall survival compared with ZFHX3 and NOTCH1/2/3 wild types and the survival benefit of the mutated and wild type genes are not significant in chemotherapy treated patients, suggesting that ZFHX3 and NTOCH2 mutations are predictive markers for guiding immune checkpoint inhibitor treatment in NSCLC [42,43]. Recent studies revealed that increased BTK expression is associated with prolonged survival in patients with lung adenocarcinoma and co-expressed genes with high expression of BTK were enriched in immune-related pathways including immune checkpoints [44] and BTK knockdown results in tumor developments and diminished CD8+ cell activity [45]. However, evidence for the association between BTK mutations and overall survival in NSCLC patients treated with immune checkpoint inhibitors remains limited. Although BTK mutation status was ranked in the PC1 loading and imputation importance in our study, BTK may be a candidate surrogate gene and may be involved in the network related to survival risk stratification in the immunotherapy treated group. This is consistent with our interpretation that the non-linear coupling pattern of 31 tumor gene mutation statuses functions as an immunotherapy-specific predictive biomarker for NSCLC patients. Several studies reported that SMARCA4 mutations are prognostic factors significantly associated with worse overall survival across treatments including with immune checkpoint inhibitors, non-immune therapies and targeted agents in NSCLC and these effects are enhanced with co-mutations in genes such as KRAS [46,47]. In the context of first-line immunotherapy vs chemoimmunotherapy in NSCLC, SMARCA4 mutation was associated with improved overall survival in immunotherapy treated patients, whereas co-mutation of SMARCA4 with NOTCH2 and ATRX was associated with worse overall survival in immunotherapy treated patients [48]. Together, these findings in the preceding studies support the biological validity of our speculation that the non-linear coupling of 31 gene mutation statuses including PIK3C2G, ZFHX3, BTK, NOTCH2 SMARCA4 captures survival heterogeneity in the immunotherapy treated patients with NSCLC.

The limitations of our study should be acknowledged. Although the clinical and gene mutation data were derived from the large real-world MSK-CHORD dataset, our data selection was limited to NSCLC tumor samples from patients treated with immune checkpoint inhibitors, either alone or in combination with chemoterapy who had recieved nivolumab receiving at least once, resulting in a small immunotherapy cohort. With no independent external validation cohort, the small sample size raises the possibility of overfitting and limiting generalizability. Further multiple validations of our DeepSurv model and the 31 gene mutation statuses are required. In addition, our model was based only on binary statuses of gene mutations. In our further work, the model will be reconstructed by incorporating other relevant clinical and molecular markers or gene expression data as input features, toward identifying regulatory networks underlying in survival risk or benefit within immuotherapy-treated patients with NSCLC.

## 4. Conclusions

In the present study, we developed a DeepSurv model, a deep learning–based extension of the Cox proportional hazards model with tumor multiple gene mutation status in NSCLC as input features to capture non-linear patterns of gene mutation combinations underlying survival heterogeneity among patients receiving immune checkpoint inhibitors including nivolumab or the immune checkpoint inhibitors including nivolumab combined with chemotherapy. We selected 31 genes as input features for the DeepSurv, based on population-level mutation frequency, patient-level variance in mutation status, and univariate Cox proportional hazards analyses evaluating the association between the presence or absence of each gene mutation and overall survival. The resulting DeepSurv-derived risk score suggested the survival heterogeneity among nivolumab-containing immunotherapy or nivolumab-containing immunotherapy combined with chemotherapy treated patients with NSCLC which was visualized as clear separation between high-risk and low-risk groups divided by the median value of the risk scores.

## Data Availability

All data produced in the present study are available upon reasonable request to the authors.

## References

[1] H. Borghaei, l. Paz-Ares, l. Horn et al. Nivolumab versus docetaxel in advanced nonsquamous non–small-cell lung cancer. N. Engl. J. Med. 2015;373, 1627–1639.

[2] Antonia SJ, Borghaei H, Ramalingam SS, et al. Four-year survival with nivolumab in patients with previously treated advanced non-small-cell lung cancer: a pooled analysis. Lancet Oncol 2019;20:1395–408.

[3] Paz-Ares LG, Ramalingam SS, Ciuleanu TE, Lee JS, Urban L, Caro RB et al. First-Line Nivolumab Plus Ipilimumab in Advanced NSCLC: 4-Year Outcomes From the Randomized, Open-Label, Phase 3 CheckMate 227 Part 1 Trial. J Thorac Oncol. 2022;17(2):289–308.

[4] Durm G, Mamdani H, Althouse S, Perkins S, Jabbour SK, Ganti AK et al. Randomized phase II study of consolidation immunotherapy with nivolumab and ipilimumab or nivolumab alone following concurrent chemoradiotherapy for unresectable stage IIIA/IIIB non-small-cell lung cancer (NSCLC): Big Ten Cancer Research Consortium LUN16-081. J Immunother Cancer. 2025;13(7):e010316.

[5] Grant MJ, Herbst RS, Goldberg SB. Selecting the optimal immunotherapy regimen in driver-negative metastatic NSCLC. Nat Rev Clin Oncol. 2021;18(10):625–644.

[6] Topalian SL, Taube JM, Anders RA, Pardoll DM. Mechanism-driven biomarkers to guide immune checkpoint blockade in cancer therapy. Nat Rev Cancer. 2016;16(5):275–87.

[7] Herbst RS, Baas P, Kim DW, Felip E, Pérez-Gracia JL, Han JY et al. Pembrolizumab versus docetaxel for previously treated, PD-L1-positive, advanced non-small-cell lung cancer (KEYNOTE-010): a randomised controlled trial. Lancet. 2016;387(10027):1540–1550.

[8] Brody R, Zhang Y, Ballas M, Siddiqui MK, Gupta P, Barker C, et al PD-L1 expression in advanced NSCLC: Insights into risk stratification and treatment selection from a systematic literature review. Lung Cancer. 2017;112:200–215.

[9] Perrone F, Leonetti A, Tiseo M, Facchinetti F. Benefit With No Target: Long-Term Outcomes of Chemoimmunotherapy in “PD-L1 Negative” NSCLC. J Thorac Oncol. 2024;19(8):1128–1132.

[10] Zhang Z, Lin Y, Chen S. Efficacy of neoadjuvant, adjuvant, and perioperative immunotherapy in non-small cell lung cancer across different PD-L1 expression levels: a systematic review and meta-analysis. Front Immunol. 2025;16:1569864.

[11] Di Federico A, Stumpo S, Mantuano F, De Giglio A, Lo Bianco F, Pecci F. et al. Long-term overall survival with dual CTLA-4 and PD-L1 or PD-1 blockade and biomarker-based subgroup analyses in patients with advanced non-small-cell lung cancer: a systematic review and reconstructed individual patient data meta-analysis. Lancet Oncol.;26(11):1443–1453.

[12] Cortellini A, Brunetti L, Di Fazio GR, Garbo E, Pinato DJ, Naidoo J. et al. Determinants of 5-year survival in patients with advanced NSCLC with PD-L1≥50% treated with first-line pembrolizumab outside of clinical trials: results from the Pembro-real 5Y global registry. J Immunother Cancer. 2025;13(2):e010674.

[13] Peters S, Paz-Ares LG, Reck M, Carbone DP, Brahmer JR, Borghaei H. et al. Long-Term Survival Outcomes With First-Line Nivolumab Plus Ipilimumab-Based Treatment in Patients With Metastatic NSCLC and Tumor Programmed Death-Ligand 1 Lower Than 1%: A Pooled Analysis. J Thorac Oncol. 2025;20(1):94–108.

[14] Rizvi NA, Hellmann MD, Snyder A, Kvistborg P, Makarov V, Havel JJ. et al. Mutational landscape determines sensitivity to PD-1 blockade in non-small cell lung cancer. Science. 2015;348(6230):124–128.

[15] Samstein RM, Lee CH, Shoushtari AN, Hellmann MD, Shen R, Janjigian YY. et al. Tumor mutational load predicts survival after immunotherapy across multiple cancer types. Nat Genet. 2019;51(2):202–206.

[16] Hellmann MD, Paz-Ares L, Bernabe Caro R, Zurawski B, Kim SW, Carcereny Costa E. et al. Nivolumab plus Ipilimumab in Advanced Non-Small-Cell Lung Cancer. N Engl J Med. 2019;381(21):2020–2031.

[17] Provencio M, Serna-Blasco R, Nadal E, Insa A, García-Campelo MR, Casal Rubio J. et al. Overall Survival and Biomarker Analysis of Neoadjuvant Nivolumab Plus Chemotherapy in Operable Stage IIIA Non-Small-Cell Lung Cancer (NADIM phase II trial). J Clin Oncol. 2022;40(25):2924–2933.

[18] Mouawad A, Boutros M, Chartouni A, Attieh F, Kourie HR. Tumor mutational burden: why is it still a controversial agnostic immunotherapy biomarker? Future Oncol. 2025;21(4):493–499.

[19] So WV, Dejardin D, Rossmann E, Charo J. Predictive biomarkers for PD-1/PD-L1 checkpoint inhibitor response in NSCLC: an analysis of clinical trial and real-world data. J Immunother Cancer. 2023;11(2):e006464.

[20] van de Haar J, Mankor JM, Hummelink K, Monkhorst K, Smit EF, Wessels LFA. et al. Combining Genomic Biomarkers to Guide Immunotherapy in Non-Small Cell Lung Cancer. Clin Cancer Res. 2024;30(7):1307–1318.

[21] Liu Z, Lin G, Yan Z, Li L, Wu X, Shi J. et al. Predictive mutation signature of immunotherapy benefits in NSCLC based on machine learning algorithms. Front Immunol. 2022;13:989275.

[22] Fomin V, So WV, Barbieri RA, Hiller-Bittrolff K, Koletou E, Tu T. et al. Machine learning identifies clinical tumor mutation landscape pathways of resistance to checkpoint inhibitor therapy in NSCLC. J Immunother Cancer. 2025;13(3):e009092.

[23] Katzman JL, Shaham U, Cloninger A, Bates J, Jiang T, Kluger Y. DeepSurv: personalized treatment recommender system using a Cox proportional hazards deep neural network. BMC Med Res Methodol. 2018;18(1):24.

[24] Jee J, Fong C, Pichotta K, Tran TN, Luthra A, Waters M., MSK Cancer Data Science Initiative Group, et al. Automated real-world data integration improves cancer outcome prediction. Nature. 2024;636(8043):728–736.

[25] Yao S, Liu X, Feng Y, Li Y, Xiao X, Han Y, Xia S. Unveiling the Role of HGF/c-Met Signaling in Non-Small Cell Lung Cancer Tumor Microenvironment. International Journal of Molecular Sciences. 2024; 25(16):9101.

[26] Sattler M, Salgia R. The expanding role of the receptor tyrosine kinase MET as a therapeutic target in non-small cell lung cancer. Cell Rep Med. 2025;6(3):101983.

[27] Cinausero M, Laprovitera N, De Maglio G, Gerratana L, Riefolo M, Macerelli M, Fiorentino M, Porcellini E, Buoro V, Gelsomino F, Squadrilli A, Fasola G, Negrini M, Tiseo M, Ferracin M, Ardizzoni A. KRAS and ERBB-family genetic alterations affect response to PD-1 inhibitors in metastatic nonsquamous NSCLC. Ther Adv Med Oncol. 2019;11:1758835919885540.

[28] Papageorgiou S, Pashley SL, O’Regan L, Khan S, Bayliss R, Fry AM. Alternative Treatment Options to ALK Inhibitor Monotherapy for EML4-ALK-Driven Lung Cancer. Cancers. 2022; 14(14):3452.

[29] Bai H, Duan J, Li C, Xie W, Fang W, Xu Y, Wang G, Wan R, Sun J, Xu J, Wang X, Fei K, Zhao Z, Cai S, Zhang L, Wang J, Wang Z. EPHA mutation as a predictor of immunotherapeutic efficacy in lung adenocarcinoma. J Immunother Cancer. 2020;8(2):e001315.

[30] Kola B, Kakkat S, Suman P, Crouch E, Chakroborty D, Sarkar C. Lymphangiogenesis in Breast Cancer: From Molecular Mechanisms to Clinical Implications. FASEB J. 2025;39(9):e70590.

[31] Langer CJ, Redman MW, Wade JL 3rd, Aggarwal C, Bradley JD, Crawford J, Stella PJ, Knapp MH, Miao J, Minichiello K, Herbst RS, Kelly K, Gandara DR, Papadimitrakopoulou VA. SWOG S1400B (NCT02785913), a Phase II Study of GDC-0032 (Taselisib) for Previously Treated PI3K-Positive Patients with Stage IV Squamous Cell Lung Cancer (Lung-MAP Sub-Study). J Thorac Oncol. 2019;14(10):1839–1846.

[32] Hayashi T, Desmeules P, Smith RS, Drilon A, Somwar R, Ladanyi M. RASA1 and NF1 are Preferentially Co-Mutated and Define A Distinct Genetic Subset of Smoking-Associated Non-Small Cell Lung Carcinomas Sensitive to MEK Inhibition. Clin Cancer Res. 2018;24(6):1436–1447.

[33] Hong D, Rasco D, Veeder M, Luke JJ, Chandler J, Balmanoukian A, George TJ, Munster P, Berlin JD, Gutierrez M, Mita A, Wakelee H, Samakoglu S, Guan S, Dimery I, Graef T, Borazanci E. A Phase 1b/2 Study of the Bruton Tyrosine Kinase Inhibitor Ibrutinib and the PD-L1 Inhibitor Durvalumab in Patients with Pretreated Solid Tumors. Oncology. 2019;97(2):102–111.

[34] Li T, Wang H, Xu J, et al. TGFBR2 mutation predicts resistance to immune checkpoint inhibitors in patients with non-small cell lung cancer. Therapeutic Advances in Medical Oncology. 2021;13.

[35] Zhang Z, Gu Y, Su X, Bai J, Guan W, Ma J, Luo J, He J, Zhang B, Geng M, Xia X, Guan Y, Shen C and Chen C. Co-Occurring Alteration of NOTCH and DDR Pathways Serves as Novel Predictor to Efficacious Immunotherapy in NSCLC. Front. Oncol. 2021;11:659321.

[36] Zheng S, Cao Y, Randall J, Yu H, Thomas TO. Integrating POLE/POLD1 mutated for immunotherapy treatment planning of advanced stage non-small cell lung cancer. Thorac Cancer. 2023;14(23):2269–2274.

[37] Czubak K, Lewandowska MA, Klonowska K, Roszkowski K, Kowalewski J, Figlerowicz M, Kozlowski P. High copy number variation of cancer-related microRNA genes and frequent amplification of DICER1 and DROSHA in lung cancer. Oncotarget. 2015;6(27):23399–416.

[38] Bai H, Zhou Y, Liu W, Xu WY, Cheng L, Huo Y, Ji H, Xiong L. Genetic mutation profiling reveals biomarkers for targeted therapy efficacy and prognosis in non-small cell lung cancer. Heliyon. 2024;10(6):e27633.

[39] Shi Y, Shin DS. Dysregulation of SWI/SNF Chromatin Remodelers in NSCLC: Its Influence on Cancer Therapies including Immunotherapy. Biomolecules. 2023; 13(6):984.

[40] Zhang, J., Zhou, N., Lin, A. et al. ZFHX3 mutation as a protective biomarker for immune checkpoint blockade in non-small cell lung cancer. Cancer Immunol Immunother 2021;70, 137–151.

[41] Gao C, Zhang J, Du X, Gao X, Diao X, Zhao K et al. Prognostic determinants and functional role of PIK3C2G in stage IIb-IIIa lung adenocarcinoma: insights from clinical and molecular analyses. Front. Oncol. 2025;14:1473437.

[42] Zhang J, Zhou N, Lin A, Luo P, Chen X, Deng H. et al. ZFHX3 mutation as a protective biomarker for immune checkpoint blockade in non-small cell lung cancer. Cancer Immunol Immunother. 2021;70(1):137–151.

[43] Zhang K, Hong X, Song Z, Xu Y, Li C, Wang G. et al. Identification of Deleterious NOTCH Mutation as Novel Predictor to Efficacious Immunotherapy in NSCLC. Clin Cancer Res. 2020;26(14):3649–3661.

[44] Jiao L, Tao Y, Ding H, Wu F, Liu Y, Li C, Li F. Bioinformatics analysis of BTK expression in lung adenocarcinoma: implications for immune infiltration, prognostic biomarkers, and therapeutic targeting. 3 Biotech. 2024;14(9):215.

[45] Huang J, Yuan Y, Guo L, Xia G, Chen Y, Chen Q, Wang M. The impact of BTK knockdown on lung adenocarcinoma growth and immune response. Cancer Sci. 2025;116(6):1550–1564.

[46] Paredes R, Borea R, Drago F, Russo A, Nigita G, Rolfo C. Genetic drivers of tumor microenvironment and immunotherapy resistance in non-small cell lung cancer: the role of KEAP1, SMARCA4, and PTEN mutations. J Immunother Cancer. 2025;13(8):e012288.

[47] Manolakos P, Boccuto L, Ivankovic DS. A Critical Review of the Impact of SMARCA4 Mutations on Survival Outcomes in Non-Small Cell Lung Cancer. J Pers Med. 2024;14(7):684.

[48] Victor Lee, Patrick Oh, Alisa Rybkin, Abhijit Patel, So Yeon Kim, and Henry Soo-Min Park Molecular markers associated with survival in PD-L1– high non-small cell lung cancer treated with first-line immunotherapy versus chemoimmunotherapy. J Clin Oncol 2025;43, e20607–e20607.

